# Sex differences in depression and substance use disorder diagnoses and overdose risk in British Columbia

**DOI:** 10.64898/2026.10.09.26364334

**Authors:** Jennifer Vincent, Heather Palis

## Abstract

**Background:** Depression is a risk factor for overdose and is more commonly diagnosed among females. However, sex-differences in the association between depression, in isolation and in combination with substance use disorder (SUD), and overdose have not been examined at the population level in British Columbia (BC). This study described sex-and-age differences in associations between documented history of depression and/or SUD diagnoses and risk of fatal (FOD) and non-fatal overdose (NFOD).

**Methods:** This study used linked administrative health data from the Provincial Health Services Authority Platform for Analytics and Data. The sample included BC residents aged 19-79 as of January 1^st^, 2015 (n=3,996,690). Exposures were classified as documented diagnosis history of 1) depression; 2) substance use disorder (SUD); 3) depression and SUD; and 4) neither condition, using physician billing and hospitalization records from 2010-2014. FOD (BC Coroners Service records) and NFOD (BC Emergency Health Services records) were ascertained from 2015 to 2024. Incidence rates per 10,000 person-years and unadjusted incidence rate ratios (IRRs) and 95% confidence intervals (CI) were estimated using Poisson regression, with neither diagnosis as reference, overall and stratified by sex and age.

**Results:** There were 14,281 FOD (78.59% male) and 43,555 NFOD (69.45% male) cases. FOD incidence was higher among males overall; however, relative associations between diagnosis history and FOD risk were stronger among females. Depression diagnosis history was associated with a FOD IRR of 3.95 (CI: 3.56-4.38) among females and 2.46 (CI: 2.32-2.61) among males. IRRs for SUD and for depression and SUD were larger among females (108.07 [CI: 95.83-121.73] and 116.70 [CI: 106.51-127.96]) than males (26.69 [CI: 25.57-28.18] and 31.21 [CI: 29.81-32.67]). NFOD findings were similar but attenuated (depression IRR: 3.22 [CI: 3.08-3.38]) in females; 2.29 [CI: 2.21-2.37] in males).

**Conclusion:** A documented history of depression and/or SUD was more strongly associated with FOD and NFOD risk among females, despite higher incidence overall among males. Future research should consider the potential role of other sociodemographic factors in explaining sex differences in associations between diagnosis history and overdose risk.

## Background

In BC and elsewhere, males have been disproportionately impacted by the unregulated drug poisoning emergency, representing the majority of both fatal and non-fatal overdoses ^1–3^. In recent years, data suggest that overdose rates in BC and the United States among females and males have diverged, with greater declines in overdose rates among males than among females ^1,2^. For instance, from 2023 to 2024, annual overdose mortality rates among males in BC declined by 15.9%, while mortality rates among females declined by 3.3%^1^. Coincidingly, sex and gender-based mental health disparities appeared to have widened since the COVID-19 pandemic, in BC and throughout Canada^4–6^. Specifically, women and females appear to have experienced greater increases in mental health conditions, with some indications that depression in particular has increased among women and females ^5,6^. Importantly, these diagnostic trends may reflect both changes in true prevalence, but also differences in patterns of care seeking and care access, which are shaped by gender alongside other interpersonal and structural factors^7,8^. Specifically, men and males are less likely to seek care for mental health concerns and are therefore less likely to receive a documented depression diagnosis ^7–9^, meaning that analyses using documented diagnosis histories in part are capturing differences in care engagement alongside differences in underlying prevalence.

People with co-occurring mental health and substance use disorder diagnoses appear to face an elevated risk of overdose, and elevated rates of discontinued substance use treatment or return to unregulated substance use while in abstinence-based substance use treatment ^10–14^ as compared to people with SUD alone. Evidence suggests that among people with SUD, concurrent mental health disorder diagnoses, particularly depression, may be more common in women and females than in men and males ^10–12,15–18^. Depression, both comorbid with substance use disorder and in isolation, is a risk factor for overdose among both sexes ^10,19,20^. While single-sex and single-gender studies have found depression to be a risk factor for overdose among women and females, this association has seldom included direct comparison to men and males^21,22^. Additionally, studies within sub-populations, such as youth, suggest that depression may have a stronger association with overdose risk among females ^23^ however these findings may not be representative of adult populations given the distinct risk factors for overdose among youth ^24,25^, and have not been conducted in BC. Furthermore, literature suggests that women and females with SUD are more likely to have comorbid depression compared to men and males with SUD ^12,18^. Nevertheless, the association of depression and SUD with overdose risk has most often been examined within the context of describing outcomes among women and females with SUD. Thus, examining depression history in isolation, as well as comorbid with SUD history, may allow for a clearer understanding of both conditions and their joint and independent contributions to overdose risk, and any sex differences therein.

Amidst the differing trends of both depression diagnoses and overdose rates by sex, and the literature indicating potential sex-differences in the association of depression and overdose among some populations (e.g. youth), there is an important knowledge gap relating to potential sex-differences in the association between depression and overdose risk, both in isolation and comorbid with SUD.

The apparent divergence in overdose rates between males and females and coinciding disproportionate increases in mental health disorder diagnoses, including depression, among females ^1–3,6,26^, indicate that there is an opportunity to better understand potential sex differences in the association between a depression diagnosis history and overdose risk at the population level. While previous studies have established depression as a risk factor for overdose ^19,21,22,24,27^, the magnitude of the effect of depression on overdose risk and how this differs by sex has not yet been adequately explored using population-level data.

### Objective

To examine the association between depression and or SUD diagnosis and overdose risk by sex.

### Hypothesis

Associations between a documented history of depression, alone and in combination with a documented history of SUD, may be larger among females than males.

## Methods

### Study Design

This was a retrospective cohort study using linked administrative health data in British Columbia, Canada. History of a documented substance use (including all substances except alcohol and tobacco) and/or depression diagnosis history within the data was ascertained between January 1, 2010, and December 31, 2014, and the overdose outcome was ascertained during a follow-up period of January 1, 2015, and December 31, 2024.

### Data sources

Data for this study was drawn from the Provincial Health Services Authority’s Platform for Analytics and Data (PANDA). PANDA contains linked administrative health data for all residents in British Columbia. These datasets are linked at the individual level using the PANDA Patient Matching Service algorithm which first applies deterministic matching using Personal Health Numbers, followed by a series of deterministic and non-deterministic matches by sex, name, and date of birth ^28^. This platform and its linkages evolved from a proof-of-concept developed during the acute phase of the COVID-19 pandemic, as described by Wilton et al. ^29,30^. To examine the association of history of a documented depression and/or substance use disorder (SUD) diagnosis history with overdose risk, and how this may differ by sex, paramedic data, hospitalization records, physician billing, and demographic information were drawn from the linked datasets listed in Table 1.

**Table 1.**
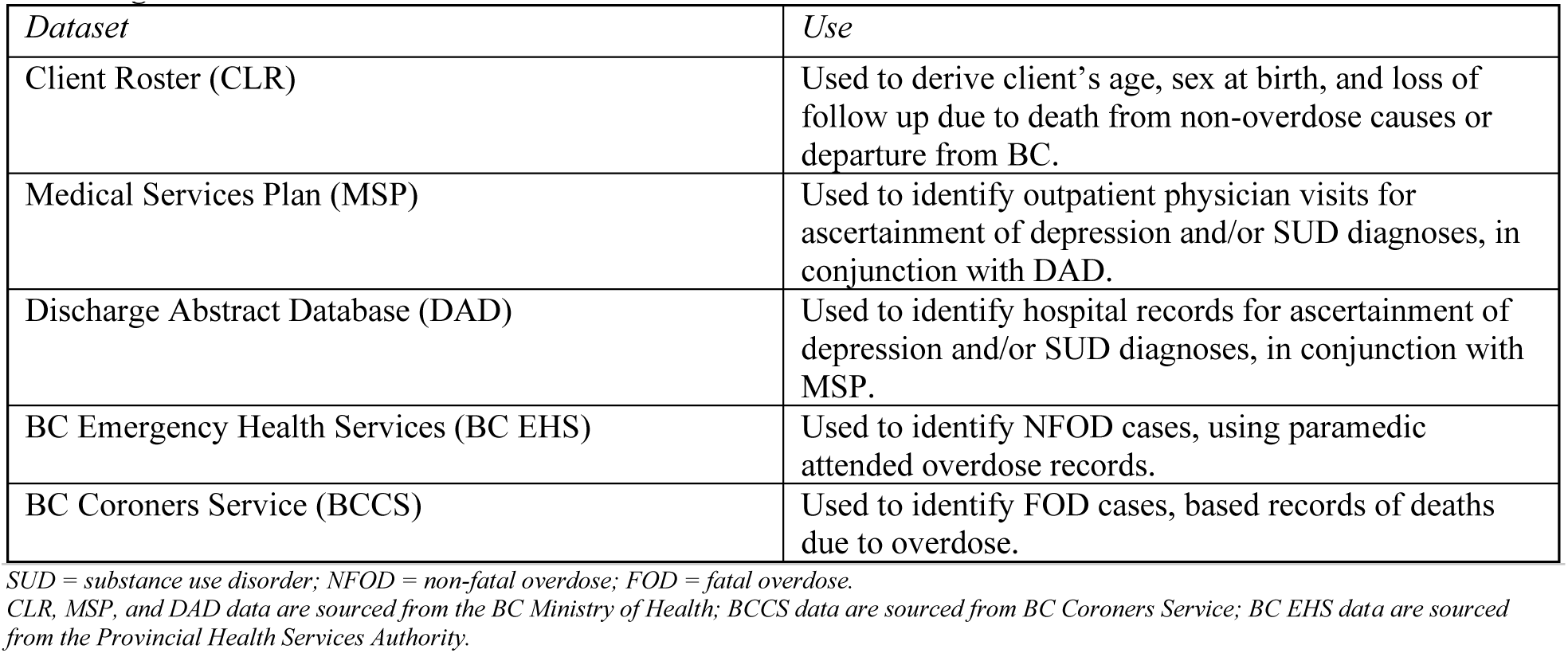
Datasets Used in the Analysis of Overdose Risk Among People with a History of a Depression and/or SUD Diagnosis in British Columbia.

| *Dataset* | *Use* |
| --- | --- |
| Client Roster (CLR) | Used to derive client's age, sex at birth, and loss of follow up due to death from non-overdose causes or departure from BC. |
| Medical Services Plan (MSP) | Used to identify outpatient physician visits for ascertainment of depression and/or SUD diagnoses, in conjunction with DAD. |
| Discharge Abstract Database (DAD) | Used to identify hospital records for ascertainment of depression and/or SUD diagnoses, in conjunction with MSP. |
| BC Emergency Health Services (BC EHS) | Used to identify NFOD cases, using paramedic attended overdose records. |
| BC Coroners Service (BCCS) | Used to identify FOD cases, based records of deaths due to overdose. |

*SUD = substance use disorder; NFOD = non-fatal overdose; FOD = fatal overdose.*
*CLR, MSP, and DAD data are sourced from the BC Ministry of Health; BCCS data are sourced from BC Coroners Service; BC EHS data are sourced from the Provincial Health Services Authority.*

Ethics approval for this study was provided by the UBC REB under approval H25-01385. All inferences, opinions, and conclusions drawn in this publication are those of the author(s), and do not reflect the opinions or policies of the Data Steward(s), including the Provincial Health Services Authority (PHSA), Ministry of Health (MoH), BC Emergency Health Services (BCEHS), and BC Coroners Service (BCCS).

### Study Sample

This study included BC residents aged 19-79 as of January 1, 2015. The study period start date reflects the earliest date for which complete overdose data were available, and the age range was selected to ensure all study members were adults at cohort entry, with age groupings used in previous BCCS reporting on overdose mortality. This sample was taken from the CLR, which contains data on BC residents with an assigned Personal Health Number (PHN)^31^. PHNs are assigned upon enrollment in the Medical Services Plan, BC’s universal health insurance plan, but may also be assigned to individuals temporarily in BC if they access healthcare while in the province. Record effective and end dates were used to identify those people actively residing in BC on January 1, 2015. Entry into the cohort was static and fixed on January 1, 2015; people who entered BC after this date were not included.

Records with unknown demographic information (age, sex) were excluded (n < 20). The true number of missing cases is likely higher than reported, owing to the patient matching algorithms inherent limitations. As PHN is only one of several factors used in matching, some contacts cannot be linked due to a missing demographic or direct identifiers (e.g. sex, name, date of birth) and would therefore not be available in the datasets to be counted; thus, it is unclear the exact number of missing cases in this study. The final study sample size of n= 3,996,690 is within 5% of the estimate of the adult BC population in BC at the midpoint of 2015 ^32,33^, indicating that the proportion of missing data is not excessive; however, these missing cases may be more likely to be structurally and/or socially marginalized ^34^, which could have a modest effect on results.

### Variables

#### Exposure

The study sample was classified into four mutually exclusive exposure groups: depression, SUD, depression and SUD, or neither. These diagnoses were derived from the MSP and DAD using the case definitions employed by the BC Chronic Disease Registry ^35–38^ and previously used in analyses relating to mental health and substance use using these data ^39,40^. Case definitions and the associated relevant ICD-9 and ICD-10 diagnostic codes (listed in Table 2) were as follows: Depression diagnosis history only

- Two or more physician visits with a depression diagnostic code at any position, OR;
- One or more hospitalizations with a depression diagnostic code at any position.

**Table 2.** ICD-9 and 10 Diagnostic Codes Used for Ascertainment of Depression and/or Substance Use Disorder Diagnosis History in British Columbia, 2010-2014.

| Diagnosis | ICD-9 Codes | ICD-10 Codes |
| --- | --- | --- |
| Depression | 296<br>311<br>50B (MSP only) | F32<br>F33 |
| SUD | 292<br>304<br>305 (Excluding 305.0 and 305.1) | F11-F16<br>F18<br>F19 |

*SUD = substance use disorder; MSP = Medical Services Plan; ICD = International Classification of Diseases. ICD-9 and ICD-10 are diagnostic coding systems used to document and classify diseases and health conditions.*

Substance use disorder diagnosis history only

- Two or more physician visits with a substance use disorder diagnostic code at any position, OR;
- One or more hospitalizations with a substance use disorder diagnostic code at any position.

Depression and substance use disorder diagnosis history

- Anyone meeting the criteria for both depression and SUD.

Neither

- Anyone not meeting the above criteria.

These criteria were applied for healthcare contacts indicating a diagnosis that occurred between January 1, 2010, and December 31, 2014. Exposure groups were mutually exclusive and fixed at the end of the ascertainment window.

While no specific validation study has been published on the depression algorithm used in this study, a similarly structured algorithm, using two physician claims or one hospitalization with a depression diagnosis within one year, showed the highest validity in a study conducted in BC and Alberta^41^. Notably, the depression algorithm used in BC includes ICD-9 code 50B, a province-specific code used for both anxiety and depression, which was not included in this validation study; the BCCDC and Ministry of Health (MoH) include this code in their algorithms due to its extensive use in the province and the presumed lack of sensitivity of analyses that do not account for this code (A. Olmstead, personal communication, 2026). Likewise, algorithms using two physician claims or one hospitalization visit with diagnostic codes indicating injection drug use within one year had the highest validity for identifying people who inject unregulated substances in prior BC and Ontario studies ^42,43^; however, these studies did not evaluate algorithm performance for people who inhale unregulated substances, who represent a growing proportion of people who use unregulated substances^44^.

#### Outcomes

##### Fatal overdose (FOD)

defined using BCCS records where the primary cause of death was determined to be due to an unintentional drug overdose^45^.

##### Non-fatal overdose (NFOD)

defined as the first paramedic-attended overdose using BCEHS call records of “Probable opioid overdose”, capturing events where naloxone was administered, paramedics assessed the overdose as being opioid-related, or 911 calls for ingestion poisoning involving paramedic-deemed cardiac arrest^46^.

FOD and NFOD were not mutually exclusive outcomes; people could be represented in both FOD and NFOD analyses if they experienced both a NFOD and FOD between January 1, 2015, and December 31, 2024.

#### Other Variables

##### Sex

Sex was defined as sex at birth, as recorded in the CLR and measured at the start of the study period.

##### Age group

Age was derived from the birth year and month in the CLR, in which the date of birth is set to the first day of the month in which the person was born, to protect privacy. Age in years was calculated as the difference between this date and the study start date, which was then classified into the following groups: 19 -29, 30-39, 40-49, 50-59, 60-69, 70-79.

### Statistical Analysis

To describe the study population, two summary tables were produced. Table 3 presents the distribution of the exposure group, sex, and age group across the full cohort and among those who experienced a fatal or non-fatal overdose during the study period. Table 4 presents the distribution of overdose outcomes, sex, and age group stratified by exposure group. Counts and column percentages are reported in both tables.

**Table 3.** Demographic and Diagnostic History Characteristics of the Full Study Cohort and Those who Experienced a Fatal or Non-Fatal Overdose in British Columbia, 2015-2024.

|  | Entire Study Cohort<br>*N* = 3,996,690<br>*N* (%) | Fatal Overdose<br>*N* = 14,281<br>*N* (%) | Non-Fatal Overdose<br>*N* = 43,555<br>*N* (%) |
| --- | --- | --- | --- |
| **DIAGNOSTIC HISTORY (2010-2014)** |  |  |  |
| Depression only | 579,729 (14.51) | 2,142 (15.00) | 7,022 (16.12) |
| Substance use disorder (SUD) only | 32,340 (0.81) | 2,140 (14.98) | 5,881 (13.50) |
| Depression and SUD | 57,566 (1.44) | 4,020 (28.15) | 11,716 (26.90) |
| Neither | 3,327,055 (83.25) | 5,979 (41.87) | 18,936 (43.48) |
| **SEX** |  |  |  |
| Male | 1,993,478 (49.88) | 11,223 (78.59) | 30,249 (69.45) |
| Female | 2,003,212 (50.12) | 3,058 (21.41) | 13,306 (30.55) |
| **AGE GROUP** |  |  |  |
| 19–29 | 846,143 (21.17) | 3,526 (24.69) | 13,937 (32.00) |
| 30–39 | 731,327 (18.30) | 3,656 (25.60) | 11,306 (25.96) |
| 40–49 | 721,998 (18.06) | 3,690 (25.84) | 8,972 (20.60) |
| 50–59 | 772,525 (19.33) | 2,826 (19.79) | 6,311 (14.49) |
| 60–69 | 614,708 (15.38) | 565 (3.96) | 2,256 (5.18) |
| 70–79 | 309,989 (7.76) | Small n | 773 (1.77) |

*SUD* = substance use disorder
Diagnostic history (depression only, SUD only, depression and SUD, neither) was ascertained from MSP data and DAD data using BC Chronic Disease Registry case definitions applied to a lookback period of January 1, 2010 – December 31, 2014. Fatal overdose was ascertained from BC Coroners Service Records; non-fatal overdose was ascertained from BC Emergency Health Services paramedic-attended overdose records. Individuals may appear in both the FOD and NFOD columns if they experienced both outcomes during follow-up.

**Table 4.** Distribution of Sex, Age Group, and Overdose Outcomes Within each Diagnostic History (2010-2014) Exposure Group Across the Full Study Cohort, British Columbia, 2015-2024.

|  | Depression only<br>*N* = 579,729<br>*N* (%) | SUD only<br>*N* = 32,340<br>*N* (%) | Depression and SUD<br>*N* = 57,566<br>*N* (%) | Neither<br>*N* = 3,327,055<br>*N* (%) |
| --- | --- | --- | --- | --- |
| **OVERDOSE OUTCOME** |  |  |  |  |
| Fatal overdose | 2,142 (0.37) | 2,140 (6.62) | 4,020 (6.98) | 5,979 (0.18) |
| *With a prior NFOD<sup>a</sup>* | 965 (45.05) | 1,173 (54.81) | 2,226 (55.37) | 2,727 (45.61) |
| Non-fatal overdose | 7,022 (1.21) | 5,881 (18.18) | 11,716 (20.35) | 18,936 (0.57) |
| *With a subsequent NFOD<sup>a</sup>* | 2,622 (37.91) | 3,274 (55.67) | 6,646 (56.73) | 7,488 (39.54) |
| **SEX** |  |  |  |  |
| Male | 199,510 (34.41) | 23,064 (71.32) | 32,573 (56.58) | 1,738,331 (52.25) |
| Female | 380,219 (65.59) | 9,276 (28.68) | 24,993 (43.42) | 1,588,724 (47.75) |
| **AGE GROUP** |  |  |  |  |
| 19–29 | 83,287 (14.37) | 6,232 (19.27) | 11,381 (19.77) | 745,243 (22.40) |
| 30–39 | 100,297 (17.30) | 8,183 (25.30) | 14,729 (25.59) | 608,118 (18.28) |
| 40–49 | 117,358 (20.24) | 7,549 (23.34) | 13,636 (23.69) | 583,455 (17.54) |
| 50–59 | 132,461 (22.85) | 6,761 (20.91) | 11,720 (20.36) | 621,583 (18.68) |
| 60–69 | 100,994 (17.42) | 2,840 (8.78) | 4,854 (8.43) | 506,020 (15.21) |
| 70–79 | 45,332 (7.82) | 775 (2.40) | 1,246 (2.16) | 262,636 (7.89) |

*SUD* = substance use disorder; *FOD* = fatal overdose; *NFOD* = non-fatal overdose
Diagnostic history (depression only, *SUD* only, depression and *SUD*, neither) was ascertained from MSP data and DAD data using BC Chronic Disease Registry case definitions applied to a lookback period of January 1, 2010 – December 31, 2014. Diagnostic groups are mutually exclusive; people meeting criteria for both depression and *SUD* are counted in the “depression and *SUD*” group only. Fatal overdose was ascertained from BC Coroners Service Records; non-fatal overdose was ascertained from BC Emergency Health Services paramedic-attended overdose records. Individuals may appear in both the *FOD* and *NFOD* columns if they experienced both outcomes during follow-up.
<sup>a</sup> Percentages for indented sub-rows are calculated among those in the parent outcome row (i.e., % of *FOD* cases with a prior *NFOD*; % of *NFOD* cases with a subsequent *FOD*), not as a proportion of the full diagnostic group column *N*.

Person-time, in years, was estimated from the CLR database, accounting for residency start and end dates, death from other causes, and the date of an individual’s FOD or first NFOD. End of residency was defined as the point at which an individual was no longer listed in the CLR as a living person in British Columbia (i.e., due to death or out-migration). Right censoring occurred at time of end of residency, non-overdose death, or the date of FOD or first NFOD. Multiple residency episodes were summed where applicable for individuals with temporary lapses in coverage within the CLR as defined above. Additionally, person-time was calculated separately for each outcome, given that the outcomes were not mutually exclusive.

Incidence rates were calculated within each diagnostic group, overall, and stratified by sex and age group. Unadjusted incidence rate ratios (IRR) and 95% confidence intervals were estimated using Poisson regression, with the “Neither” group as the reference category. Models were run overall, stratified by sex, and stratified by age, overall and by sex. The log of total person time was used as the offset in these Poisson models.

Given the temporal variation in overdose rates driven by changes in the unregulated drug supply ^47,48^ and the effects of the COVID-19 pandemic ^49^, year-adjusted IRRs were estimated to assess whether the year of the overdose had any association with the diagnostic group estimates. Poisson regression models with the yearly overdose counts as the outcome and the year the overdose occurred included as a categorical variable, with 2015 as the reference year, as it was the last full calendar year prior to the declaration of a public health emergency for the unregulated drug poisoning emergency in BC ^50^. Person-time was calculated using the same method described above but partitioned by calendar year rather than aggregated across the full study period; the log of each year’s person-time was used as the offset in these models to account for year as a time-varying variable. These models, and their stratifications, were run separately for FOD and NFOD. All analyses were performed in R (version 4.3.1).

## Results

### Cohort Characteristics

#### Overall Cohort and Diagnostic Group Distribution

Table 3 presents the demographic and diagnostic history characteristics of the full study cohort (n = 3,996,690) as well as those who experienced a fatal (n=14,281) or non-fatal (n=43,555) overdose during the study period. Note that these groups are not mutually exclusive; individuals who experienced a first NFOD prior to a FOD may appear in both outcome groups.

Though those with a history of depression and SUD diagnoses comprised 1.44% of the overall cohort, and those with a history of SUD diagnosis alone 0.81%, these groups accounted for nearly one-third and one-sixth of FOD (28.15%, 14.98%) and NFOD (26.90%, 13.50%) cases, respectively. Notably, depression and SUD diagnosis history was more prevalent than SUD diagnosis history alone across all groups, reflecting a high co-occurrence of depression with SUD in BC. Depression diagnosis history (14.51% overall) was marginally more common among those with an NFOD (16.12%) or FOD (15.00%) than the overall population. The ‘neither’ group was, correspondingly, underrepresented among NFOD (43.48%) and FOD (41.87%) cases compared to the overall cohort (83.25%).

Males comprised 49.88% of the overall cohort and a disproportionately higher proportion of both FOD (78.59%) and NFOD (69.45%) cases. Notably, females represented a relatively higher proportion of NFOD (30.55%) than FOD (21.41%) cases.

FOD cases tended to be slightly older than NFOD cases. For example, the 19-29 age group accounted for 24.69% of FOD and 32.00% of NFOD cases. Meanwhile, the 40-49 and 50-59 age groups accounted for 25.84% and 19.79% of all FOD cases, respectively, compared to 20.60% and 14.49% of NFOD cases.

#### Cohort Characteristics by Diagnostic Group

Table 4 presents the demographic characteristics and overdose outcomes of the cohort stratified by exposure group.

The depression and SUD diagnosis history and SUD diagnosis history groups had the highest proportions of fatal and non-fatal overdoses, while the neither group had the lowest. FOD and NFOD occurred in 0.37% and 1.21% of those with a depression diagnosis history, 6.62% and 18.18% of those with a SUD diagnosis history, 6.98% and 20.35% of those with a depression and SUD diagnosis history, and 0.18% and 0.57% of those with a history of neither condition. This pattern was similar for NFOD occurring prior to FOD, where among those in the SUD and depression and SUD history groups 54.81% and 55.37% who experienced FOD had a prior NFOD, compared to 45.05% and 45.61% in the depression history and neither groups. Likewise, 55.67% and 56.73% in the SUD and depression and SUD history groups experienced subsequent NFOD after their initial NFOD, compared to 37.91% and 39.54% of the depression history and neither groups.

The 19-29 age group was underrepresented across all diagnostic history groups. This was particularly pronounced for the depression history group, where 14.37% of the depression history group was 19-29, compared to 22.40% of the neither group. The depression and SUD history and SUD history groups had similar age profiles to one another, and similarly differed from the depression history and neither group. Specifically, the 30-39 (depression and SUD: 25.59%, SUD: 25.30%) and 40-49 (depression and SUD: 23.69%, SUD: 23.34%) groups were proportionally larger than in the neither group (30-39: 18.28%, 40-49: 17.54%) and depression diagnosis history group (30-39: 17.30%, 40-49: 20.24%). Accordingly, the 60-69 (depression and SUD: 8.43%, SUD: 8.78%) and 70-79 (depression and SUD: 2.40%, SUD: 2.16%) age groups were proportionally smaller than in the neither (60-69:15.21%, 70-79: 7.89%) and depression diagnosis history (60-60: 17.42%, 70-79: 7.82%) groups.

Depression diagnosis history was more common among females (65.59%) than males (34.41%), while SUD diagnosis history was more common among males (71.32%) than females (28.68%); the depression and SUD diagnosis history group was somewhat more evenly split, with males comprising 56.58% and females comprising 43.42%.

### Incidence Rates

Table 5 and Table 6 present overall and age- and sex-stratified overdose incidence rates per 10,000 person-years and unadjusted IRRs for FOD and NFOD, respectively. Overall, the incidence of FOD was 3.92 per 10,000 person-years among people with a depression diagnosis history, 75.61 per 10,000 person-years among people with a SUD diagnosis history, 80.63 per 10,000 person-years among people with a depression and SUD diagnosis history, and 1.89 per 10,000 person-years among people with neither condition. Corresponding NFOD incidence rates were 12.88, 222.84, 252.31, and 5.99 per 10,000 person-years. Among those with neither condition, incidence of NFOD was highest in the 19-29 group and declined with age, overall and in sex-stratified analyses. For FOD, incidence for people with a history of neither condition was highest in the 19-29 group and declined with age for the overall and female strata, while for males incidence was highest in the 30-39 group, after which it declined with age.

**Table 5.**
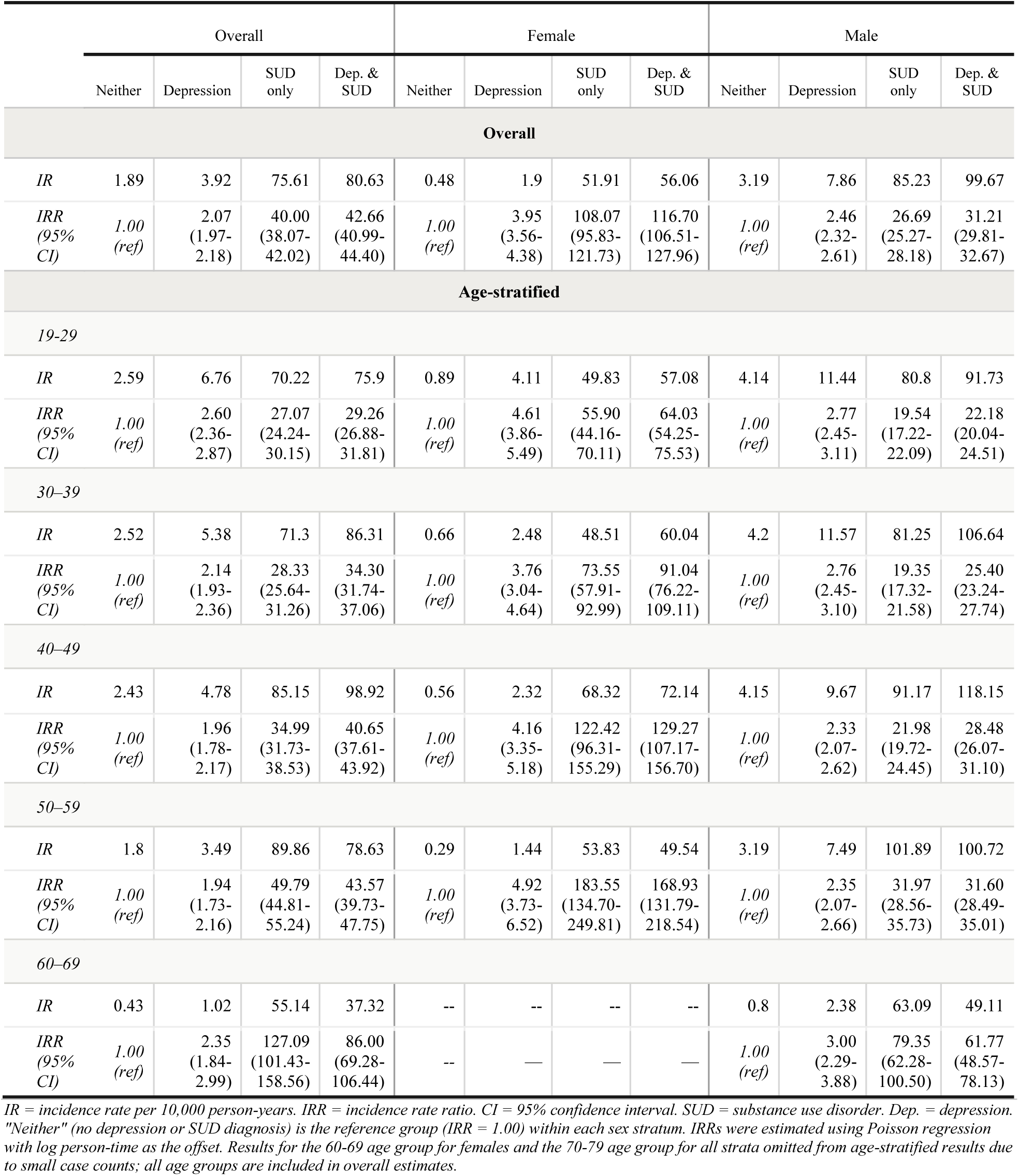
Unadjusted Incidence Rates and Incidence Rate Ratios for Fatal Overdose by Diagnostic History Group, overall and stratified by sex and age group, British Columbia, 2015-2024.

|  | Overall |  |  |  | Female |  |  |  | Male |  |  |  |
| --- | --- | --- | --- | --- | --- | --- | --- | --- | --- | --- | --- | --- |
|  | Neither | Depression | SUD only | Dep. & SUD | Neither | Depression | SUD only | Dep. & SUD | Neither | Depression | SUD only | Dep. & SUD |
| **Overall** |  |  |  |  |  |  |  |  |  |  |  |  |
| *IR* | 1.89 | 3.92 | 75.61 | 80.63 | 0.48 | 1.9 | 51.91 | 56.06 | 3.19 | 7.86 | 85.23 | 99.67 |
| *IRR (95% CI)* | *1.00 (ref)* | 2.07 (1.97-2.18) | 40.00 (38.07-42.02) | 42.66 (40.99-44.40) | *1.00 (ref)* | 3.95 (3.56-4.38) | 108.07 (95.83-121.73) | 116.70 (106.51-127.96) | *1.00 (ref)* | 2.46 (2.32-2.61) | 26.69 (25.27-28.18) | 31.21 (29.81-32.67) |
| **Age-stratified** |  |  |  |  |  |  |  |  |  |  |  |  |
| *19-29* |  |  |  |  |  |  |  |  |  |  |  |  |
| *IR* | 2.59 | 6.76 | 70.22 | 75.9 | 0.89 | 4.11 | 49.83 | 57.08 | 4.14 | 11.44 | 80.8 | 91.73 |
| *IRR (95% CI)* | *1.00 (ref)* | 2.60 (2.36-2.87) | 27.07 (24.24-30.15) | 29.26 (26.88-31.81) | *1.00 (ref)* | 4.61 (3.86-5.49) | 55.90 (44.16-70.11) | 64.03 (54.25-75.53) | *1.00 (ref)* | 2.77 (2.45-3.11) | 19.54 (17.22-22.09) | 22.18 (20.04-24.51) |
| *30-39* |  |  |  |  |  |  |  |  |  |  |  |  |
| *IR* | 2.52 | 5.38 | 71.3 | 86.31 | 0.66 | 2.48 | 48.51 | 60.04 | 4.2 | 11.57 | 81.25 | 106.64 |
| *IRR (95% CI)* | *1.00 (ref)* | 2.14 (1.93-2.36) | 28.33 (25.64-31.26) | 34.30 (31.74-37.06) | *1.00 (ref)* | 3.76 (3.04-4.64) | 73.55 (57.91-92.99) | 91.04 (76.22-109.11) | *1.00 (ref)* | 2.76 (2.45-3.10) | 19.35 (17.32-21.58) | 25.40 (23.24-27.74) |
| *40-49* |  |  |  |  |  |  |  |  |  |  |  |  |
| *IR* | 2.43 | 4.78 | 85.15 | 98.92 | 0.56 | 2.32 | 68.32 | 72.14 | 4.15 | 9.67 | 91.17 | 118.15 |
| *IRR (95% CI)* | *1.00 (ref)* | 1.96 (1.78-2.17) | 34.99 (31.73-38.53) | 40.65 (37.61-43.92) | *1.00 (ref)* | 4.16 (3.35-5.18) | 122.42 (96.31-155.29) | 129.27 (107.17-156.70) | *1.00 (ref)* | 2.33 (2.07-2.62) | 21.98 (19.72-24.45) | 28.48 (26.07-31.10) |
| *50-59* |  |  |  |  |  |  |  |  |  |  |  |  |
| *IR* | 1.8 | 3.49 | 89.86 | 78.63 | 0.29 | 1.44 | 53.83 | 49.54 | 3.19 | 7.49 | 101.89 | 100.72 |
| *IRR (95% CI)* | *1.00 (ref)* | 1.94 (1.73-2.16) | 49.79 (44.81-55.24) | 43.57 (39.73-47.75) | *1.00 (ref)* | 4.92 (3.73-6.52) | 183.55 (134.70-249.81) | 168.93 (131.79-218.54) | *1.00 (ref)* | 2.35 (2.07-2.66) | 31.97 (28.56-35.73) | 31.60 (28.49-35.01) |
| *60-69* |  |  |  |  |  |  |  |  |  |  |  |  |
| *IR* | 0.43 | 1.02 | 55.14 | 37.32 | -- | -- | -- | -- | 0.8 | 2.38 | 63.09 | 49.11 |
| *IRR (95% CI)* | *1.00 (ref)* | 2.35 (1.84-2.99) | 127.09 (101.43-158.56) | 86.00 (69.28-106.44) | -- | — | — | — | *1.00 (ref)* | 3.00 (2.29-3.88) | 79.35 (62.28-100.50) | 61.77 (48.57-78.13) |

*IR* = incidence rate per 10,000 person-years. *IRR* = incidence rate ratio. *CI* = 95% confidence interval. *SUD* = substance use disorder. *Dep.* = depression. "Neither" (no depression or SUD diagnosis) is the reference group (*IRR* = 1.00) within each sex stratum. *IRRs* were estimated using Poisson regression with log person-time as the offset. Results for the 60-69 age group for females and the 70-79 age group for all strata omitted from age-stratified results due to small case counts; all age groups are included in overall estimates.

**Table 6.**
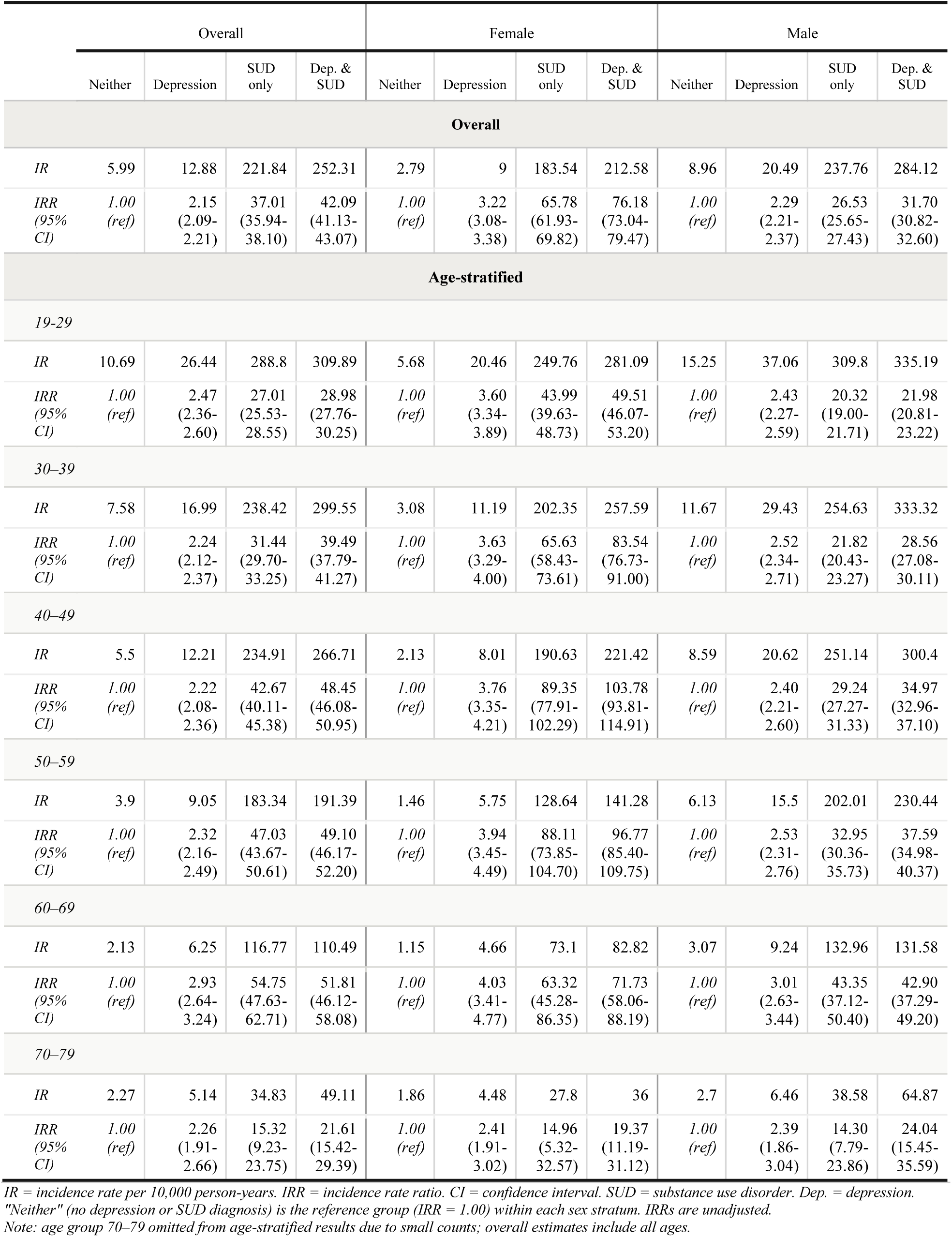
Unadjusted Incidence Rates and Incidence Rate Ratios for Non-Fatal Overdose by Diagnostic Group, Overall and Stratified by Sex and Age Group, British Columbia, 2015-2024.

Incidence was higher among males across all diagnostic groups and both outcomes. Among males, FOD incidence was 7.86 per 10,000 person-years for depression diagnosis history, 85.23 per 10,000 person-years for SUD diagnosis history, 99.67 for depression and SUD diagnosis history, and 3.19 for neither condition, respectively. Among females, the corresponding FOD incidence rates were 1.90, 51.91, 56.06, and 0.48 per 10,000 person-years. For NFOD, the incidence among males was 20.49 per 10,000 person-years, 237.76 per 10,000 person-years, 284.12 per 10,000 person-years, and 8.96 per 10,000 person-years for those with a depression diagnosis history, SUD diagnosis history, depression and SUD diagnosis history, and neither condition, respectively; for females, these rates were 9.00, 183.54, 212.58, and 2.79 per 10,000 person-years. This pattern of males having higher overdose incidence than females persisted with age stratification, for example, in the 19-29 age group with neither condition, FOD incidence was 4.14 per 10,000 person-years in males and 0.89 per 10,000 person-years in females.

### Incidence Rate Ratios – Fatal Overdose

#### Incidence Rate Ratios by Diagnostic History, Sex, and Age

Despite higher absolute incidence rates among males, the relative effects of each diagnostic history group were larger among females. Overall, depression diagnosis history, SUD diagnosis history, and depression and SUD diagnosis history were associated with 2.07 (CI: 1.97-2.18), 40.00 (CI: 38.07-42.66), and 42.66 (CI: 40.99-44.40) times the FOD risk relative to people with neither condition. The effects were similar in direction among males (depression IRR (CI): 2.46 (2.32-2.61), SUD IRR (CI): 26.69 (25.27-28.18), depression and SUD IRR (CI): 31.21 (29.81 - 32.67)) and females (depression IRR (CI): 3.95 (3.56-4.38), SUD IRR (CI): 108.07 (95.83-121.73), depression and SUD IRR (CI): 116.70 (106.51-127.96)), indicating that overall and across all sex strata, depression and SUD and SUD diagnosis histories were associated with the greatest relative increases in FOD risk, with a depression diagnosis history contributing to significant risk as well. Notably, the association between a depression diagnosis history and overdose risk relative to neither group was weaker in the overall analysis than in either sex strata. Age-stratified overall IRRs were broadly consistent with these patterns, with the attenuation of the depression IRR in the overall estimates relative to both males and females present across all age groups. As well, in overall analyses the association between depression and FOD risk appears highest in the 19-29 and 60-69 age groups, while the associations between depression and SUD and SUD diagnosis history and overdose risk appear to increase with age (Figure 1).

**Figure 1.**
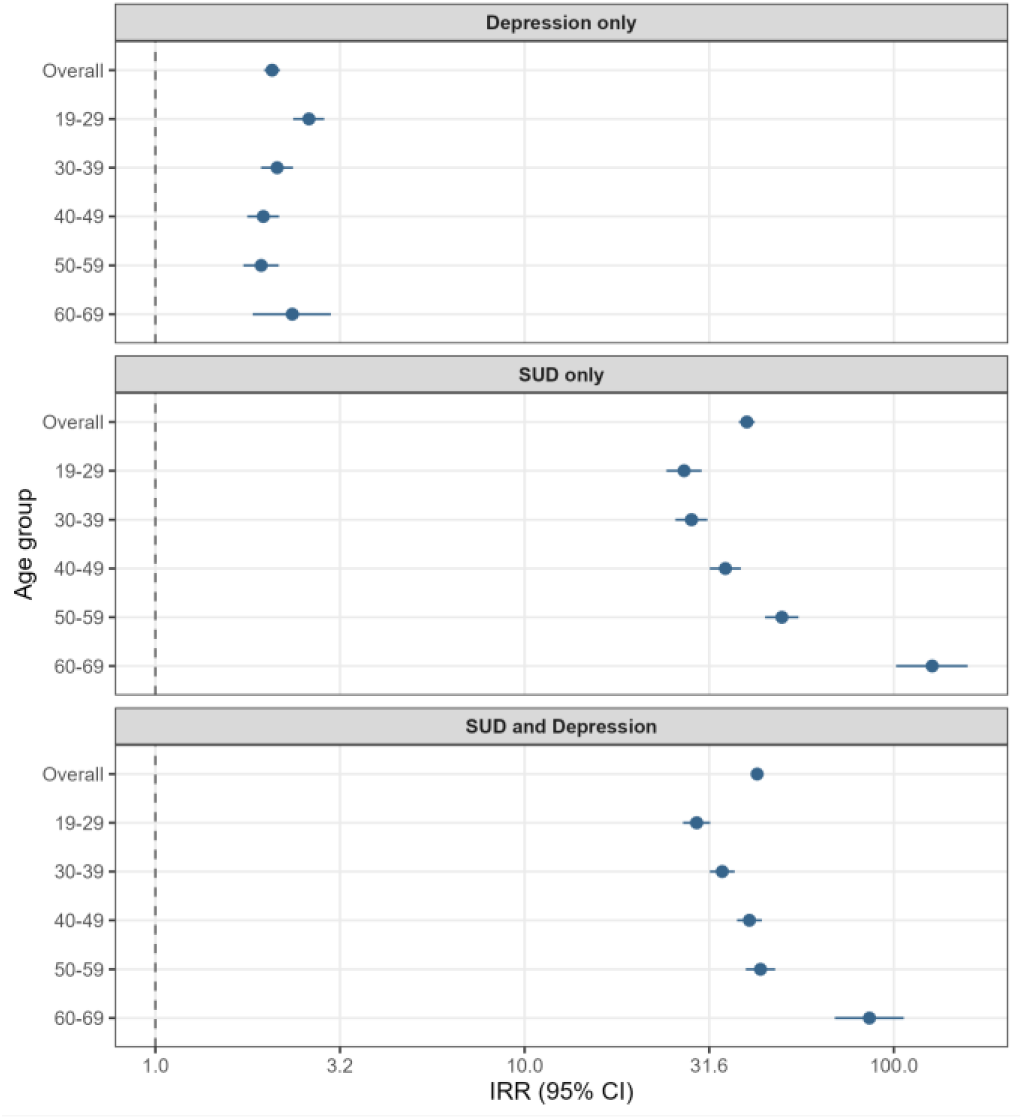
Unadjusted Incidence Rate Ratios and 95% Confidence Intervals for Fatal Overdose by Diagnostic History Group, Relative to the Neither Group, Overall and Stratified by Age, British Columbia, 2015-2024.

##### Age-stratified Incidence Rate Ratios – Males

IRRs for FOD among males were relatively stable across the 19-29 to 40-49 age groups, with depression and SUD diagnosis history having the largest effect on FOD risk, closely followed by SUD diagnosis history (Figure 2). In the 50-59 age group, the IRR for SUD diagnosis history (IRR: 31.97, CI:28.56 - 35.73) is roughly equivalent to that for depression and SUD diagnosis history (IRR: 31.60, CI: 28.59 - 35.01). In the 60-69 age group, the IRR for SUD diagnosis history of IRR: 79.35 (CI: 62.28 - 100.50) exceeded that of depression and SUD diagnosis history (IRR: 61.77, CI: 48.57 - 78.13), though CIs overlapped. The elevated FOD incidence in the neither group and the underrepresentation in all diagnostic groups of the 19-29 group may be compressing the IRR estimates for this age group, potentially understating the associations between depression and/or SUD diagnoses and overdose risk among younger individuals.

**Figure 2.**
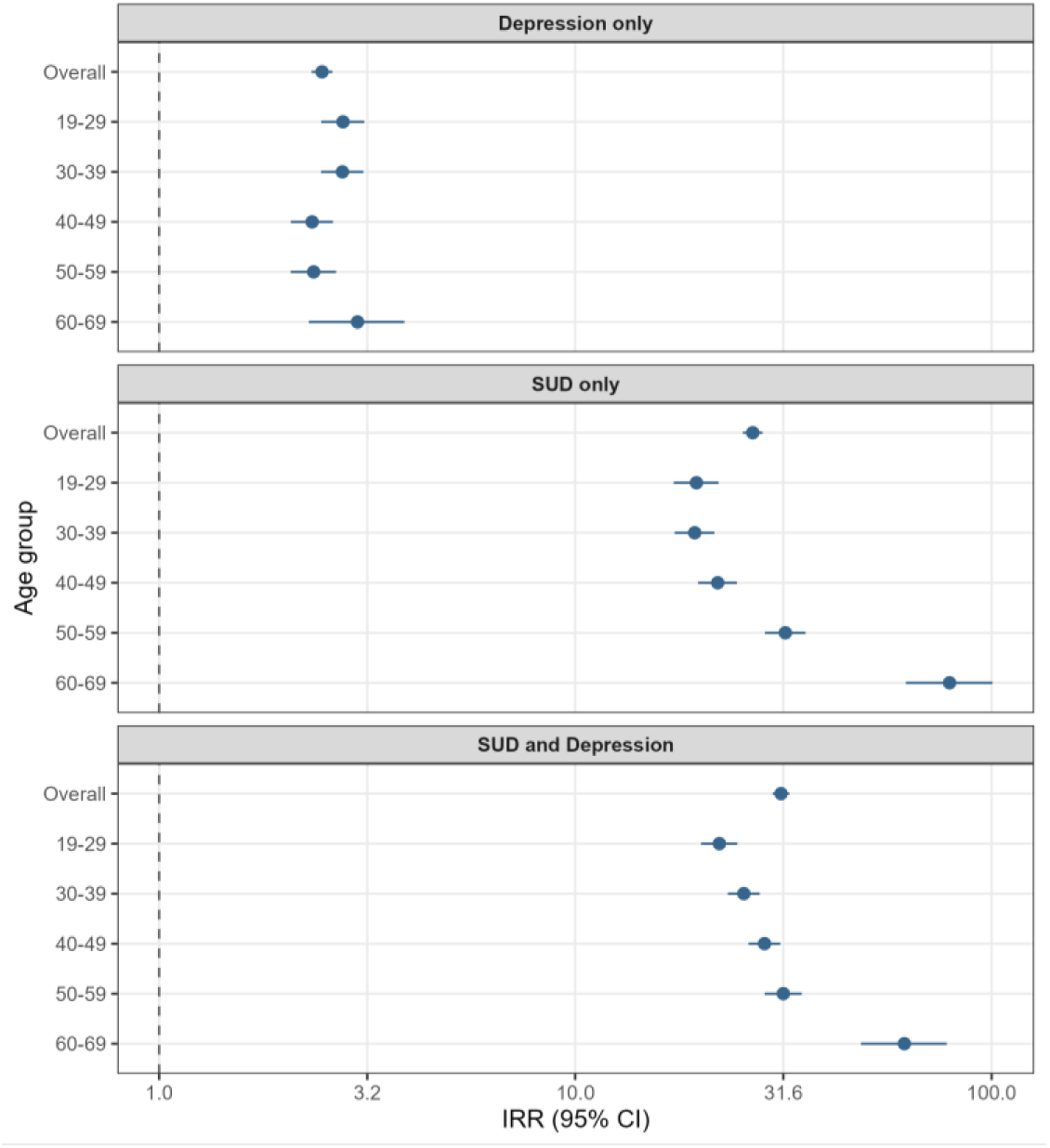
Unadjusted Incidence Rate Ratios and 95% Confidence Intervals for Fatal Overdose by Diagnostic History Group, Relative to the Neither Group, Overall and Stratified by Age, Among Males, British Columbia, 2015-2024.

##### Age-stratified Incidence Rate Ratios– Females

Among females, the associations between different diagnostic histories and overdose risk appeared more varied with age. Notably, the IRR for depression diagnosis history followed a non-linear pattern across age groups among females, with the IRR declining from the 19-29 group (IRR: 4.61, CI: 3.86-5.49) to the 30-39 group (IRR: 3.76, CI: 3.04 - 4.64) before increasing again with age (Figure 3).

**Figure 3.**
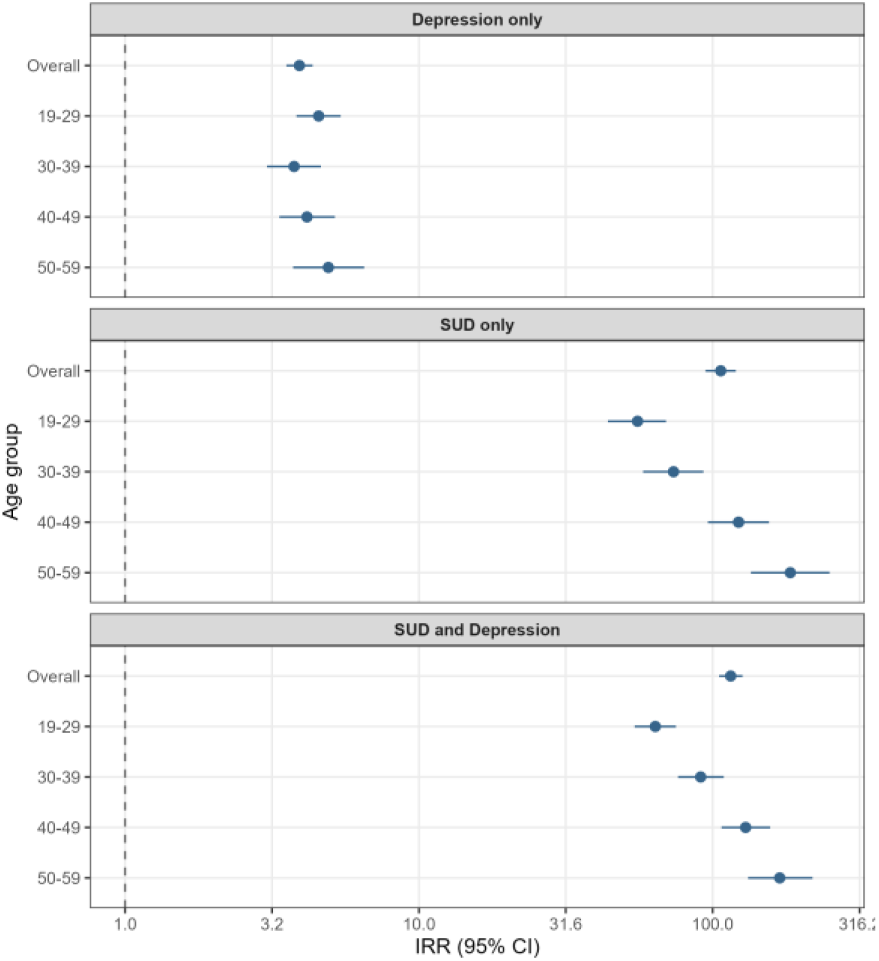
: Unadjusted Incidence Rate Ratios and 95% Confidence Intervals for Fatal Overdose by Diagnostic History Group, Relative to the Neither Group, Overall and Stratified by Age, Among Females, British Columbia, 2015-2024.

Depression and SUD and SUD diagnosis history IRRs increased with age, with the IRR estimates for depression and SUD and SUD diagnosis history in the 50-59 age group being roughly three times those seen in the 19-29 age group, in contrast to the relative stability seen in males. As with males, the SUD diagnosis history IRR eventually exceeded that of depression and SUD diagnosis history by the 50-59 group.

### Incidence Rate Ratios – Non-Fatal Overdose

#### Overall Non-Fatal Overdose Incidence Rate Ratios

NFOD IRR patterns mirrored those of FOD, with the caveat that the effect sizes for all conditions were attenuated overall and for females relative to FOD, and to a lesser extent for depression diagnosis history effect sizes among males. Overall, depression, SUD, and depression and SUD diagnosis history were responsible for 2.15 (CI: 2.09-2.21), 37.01 (CI: 35.94-38.10), and 42.09 (CI: 41.13-43.07) times the NFOD risk, respectively. Among males, IRRs were 2.29 (CI: 2.21-2.37) for depression diagnosis history, 26.53 (CI: 25.65 - 27.43) for SUD diagnosis history, and 31.71 (CI: 30.82 - 32.60) for depression and SUD diagnosis history. Among females, IRRs were 3.22 (CI: 3.08 - 3.38) for depression diagnosis history, 65.78 (CI: 61.93 - 69.82) for SUD diagnosis history, and 76.18 (CI: 73.04 - 79.47) for depression and SUD diagnosis history. As with analysis of FOD risk, the association between depression and NFOD risk appears strongest in the 19-29 and 60-69 age groups (Figure 4). As well, the associations between depression and SUD and SUD diagnosis histories and NFOD risk also appear to increase with age until the 70-79 age group, at which point they decrease—noting that a direct comparison to FOD risk in this instance is not possible due to cell count suppression.

**Figure 4.**
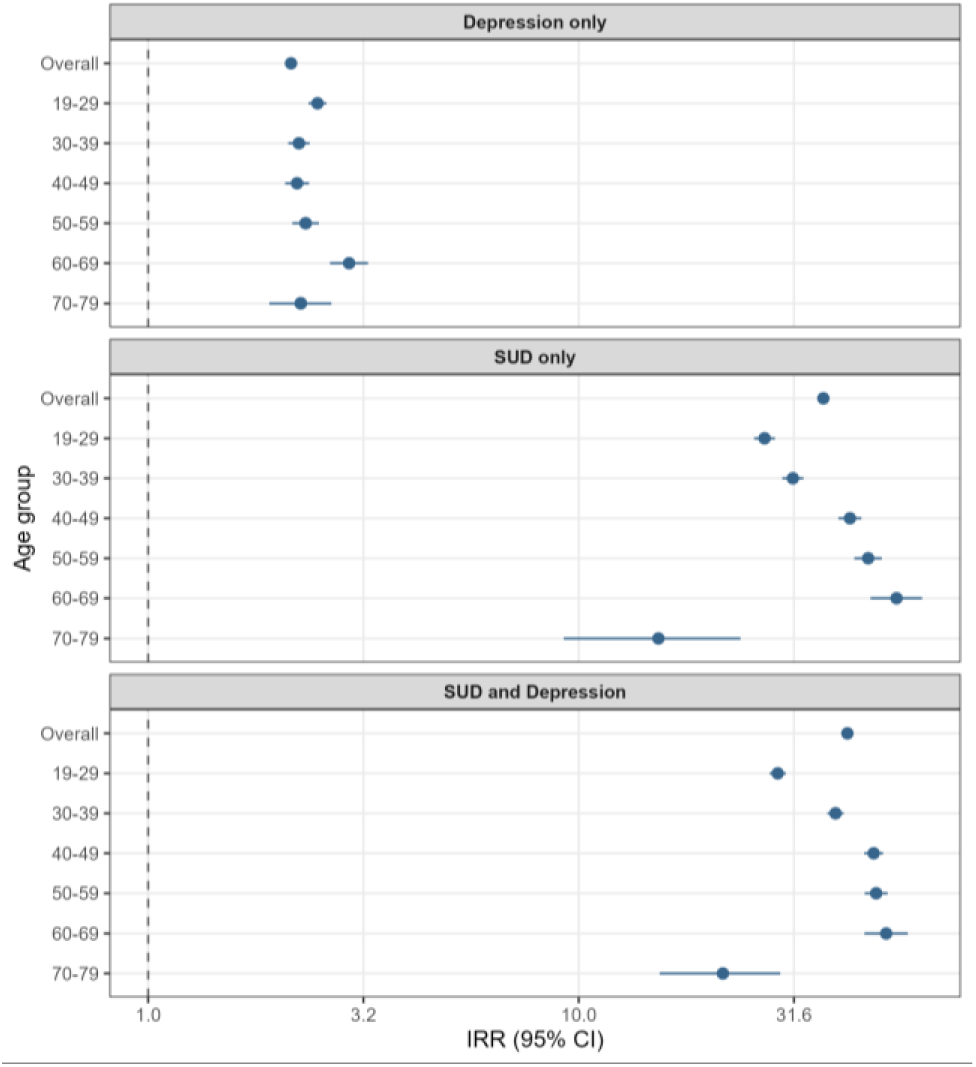
Unadjusted Incidence Rate Ratios and 95% Confidence Intervals for Non-Fatal Overdose by Diagnostic History Group, Relative to the Neither Group, Overall and Stratified by Age, British Columbia, 2015-2024.

##### Age-stratified Incidence Rate Ratios – Males

As with FOD, IRRs remained relatively stable across age groups among males, with attenuation of depression diagnosis history effect sizes relative to FOD (Figure 5). The weakest associations were seen in the 19-29 group, potentially reflecting the compression by elevated baseline incidence in the neither group in this age group.

**Figure 5.**
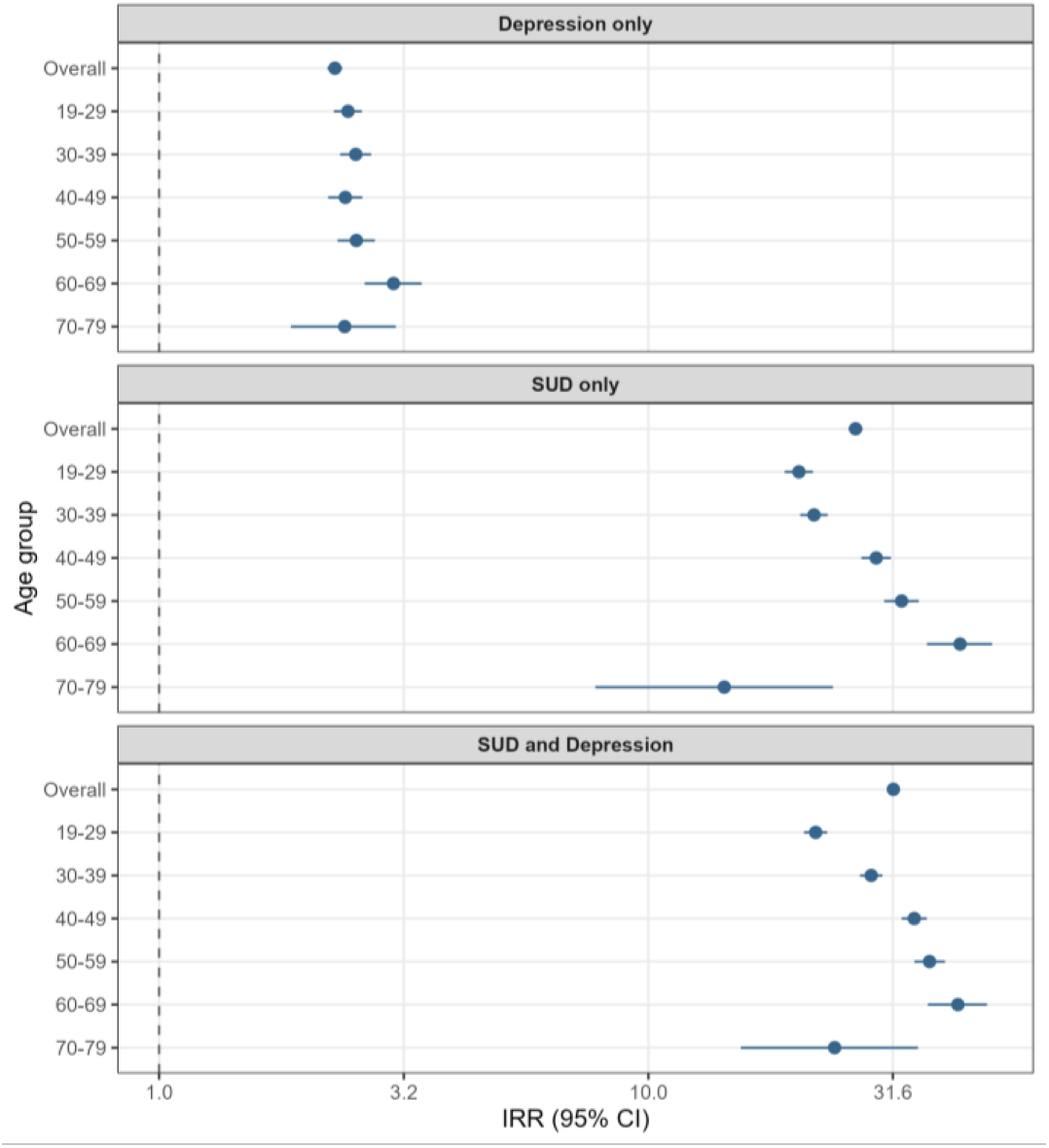
Unadjusted Incidence Rate Ratios and 95% Confidence Intervals for Non-Fatal Overdose by Diagnostic History Group, Relative to the Neither Group, Overall and Stratified by Age, Among Males, British Columbia, 2015-2024.

##### Age-stratified Incidence Rate Ratios – Females

As with males, effect sizes for NFOD were consistently attenuated relative to FOD among females (Figure 6). In contrast to the non-linear pattern observed for FOD in Section 2.3.3.3, the effect of depression diagnosis history on NFOD risk increases more uniformly with age among females, which may point to differing mechanisms operating across the two outcomes, particularly at younger ages.

**Figure 6.**
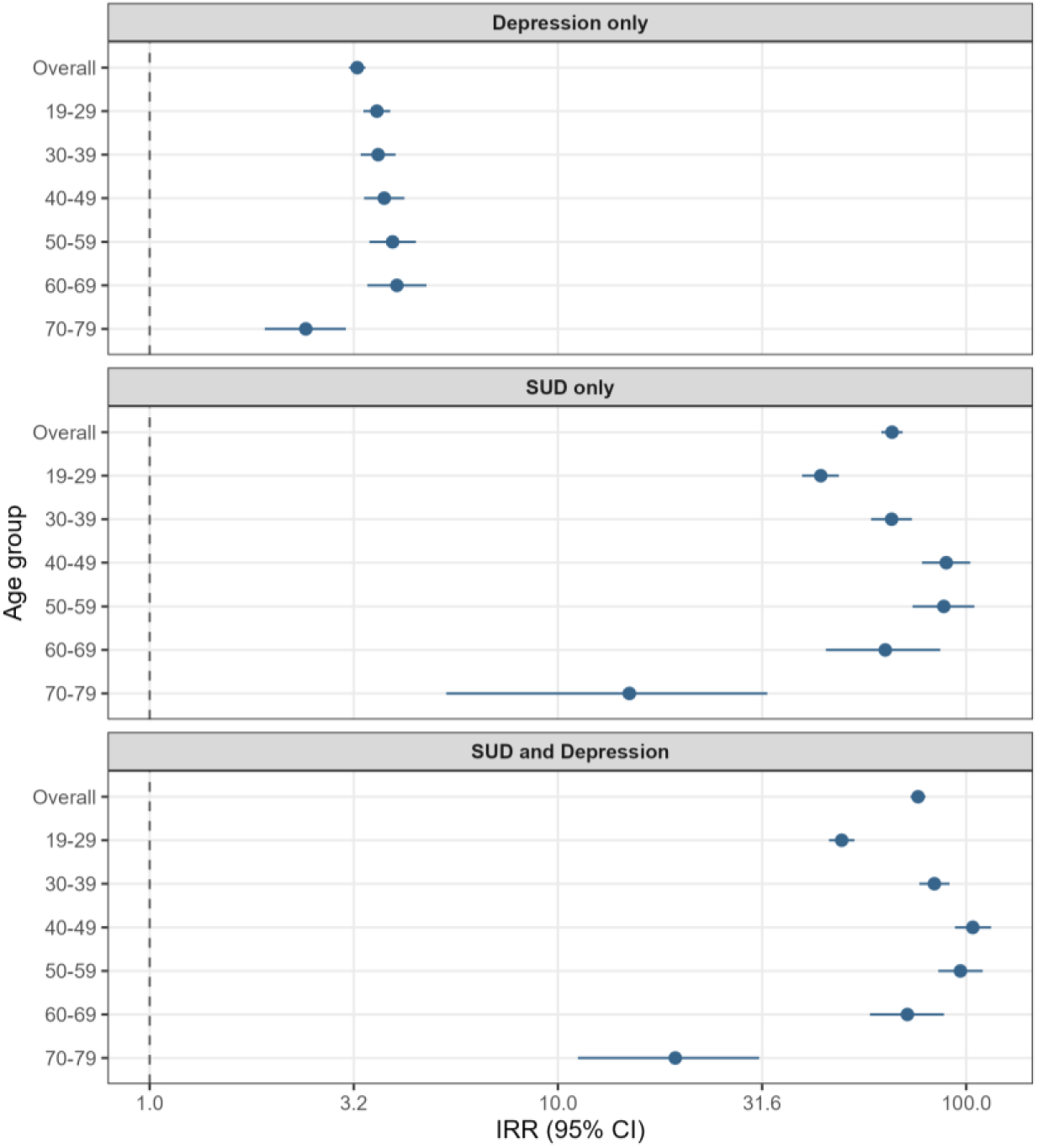
Unadjusted Incidence Rate Ratios and 95% Confidence Intervals for Non-Fatal Overdose by Diagnostic History Group, Relative to the Neither Group, Overall and Stratified by Age, Among Females British Columbia, 2015-2024.

###### Year - Adjusted Incidence Rate Ratio Analysis

Adjusting for year did not meaningfully alter effect estimates for any of the diagnostic groups for either outcome, across any strata (Appendices A.1 and A.2). This suggests that the unregulated drug poisoning emergency has affected people in these diagnostic groups roughly proportionally to their underlying risk over time, and reinforces the robustness and strength of SUD, and to a lesser extent depression, as risk factors for FOD and NFOD.

### Sex-stratified Comparison Using Depression Diagnosis History as the Reference Group

Table 7 presents unadjusted IRRs for FOD and NFOD respectively, using the depression diagnosis history group as the reference, stratified by sex.

**Table 7.** Unadjusted Incidence Rate Ratios for Fatal and Non-Fatal Overdose by Diagnostic Group and Sex, Using the Depression-Only Group as Reference, British Columbia, 2015-2024.

| Outcome | Overall |  |  | Female |  |  | Male |  |  |
| --- | --- | --- | --- | --- | --- | --- | --- | --- | --- |
|  | Depression | SUD and Depression | SUD only | Depression | SUD and Depression | SUD only | Depression | SUD and Depression | SUD only |
| FOD —<br>IRR<br>(95% CI) | 1.00 (ref) | 20.58<br>(19.53-21.69) | 19.30<br>(18.18-20.49) | 1.00 (ref) | 29.55<br>(26.92-32.46) | 27.36<br>(24.23-30.86) | 1.00 (ref) | 12.68<br>(11.90-13.51) | 10.84<br>(10.11-11.63) |
| NFOD<br>— IRR<br>(95% CI) | 1.00 (ref) | 19.59<br>(19.02-20.18) | 17.22<br>(16.64-17.83) | 1.00 (ref) | 23.63<br>(22.58-24.72) | 20.40<br>(19.16-21.70) | 1.00 (ref) | 13.87<br>(13.34-14.43) | 11.61<br>(11.11-12.12) |

IRR = unadjusted incidence rate ratio. CI = 95% confidence interval. FOD = fatal overdose. NFOD = non-fatal overdose. SUD = substance use disorder. The “depression only” group is the reference category (IRR = 1.00) within each sex stratum. IRRs were estimated using Poisson regression with log person-time as the offset.

The step-up in risk from depression diagnosis history to SUD or depression and SUD diagnosis history is larger among females than males for both FOD and NFOD. For FOD, the IRRs for depression and SUD and SUD diagnosis histories among females were 29.55 (CI: 26.92-32.46) and 27.36 (CI: 24.23-30.86), respectively, while the corresponding IRRs for males were 10.84 (CI: 10.11-11.63) and 12.65 (CI: 11.88-13.48). Further, as in Section 2.3.4.1, effect sizes were larger among females for FOD compared to NFOD, where the IRRs for depression and SUD and SUD diagnosis history among females were 20.09 (CI: 18.88-21.37) and 23.28 (CI: 22.25-24.35), respectively; this pattern was not seen among males.

## Discussion

The substantially higher prevalence of a documented depression and/or SUD diagnosis history compared to a SUD diagnosis history in isolation seen in the descriptive statistics aligned with past research on mental health and overdose in BC^51^. The relatively higher proportion of females in non-fatal overdose cases compared to fatal overdose cases was also consistent with prior analyses of overdose in BC ^49,52^. Documented diagnosis estimates were similar to previous studies in the province, with the 5-year estimates used here between BC surveillance figures for lifetime prevalence and annual incidence for both conditions within the study’s time period ^53^, as expected given the intermediate lookback window used.

The greater proportion of females among documented depression diagnosis cases is unsurprising given the higher prevalence of depression among females ^4,9,54^ and underdiagnosis of depression among males ^7^. This underdiagnosis is particularly relevant to the interpretation of findings for the “neither” group among males, insofar as those without a documented diagnosis of depression or SUD may not necessarily represent a wholly healthier population, so much as one that has less engagement with healthcare. Similarly, the relative sex parity of the depression and SUD group, in contrast to the substantially higher proportion of males with SUD alone, may reflect the increased likelihood of females to have a depression disorder diagnosis comorbid with SUD ^10^.

The lower overall overdose incidence but greater associations between diagnostic history and overdose risk seen in females may reflect a concentration of overdose risk among diagnosed females, whereas among males overdose risk may be more broadly distributed. Additionally, the greater effect sizes for FOD versus NFOD for females, and the larger step-up in risk between depression diagnosis history and depression and SUD or SUD diagnosis history seen among females may further suggest that overdose risk is more sharply concentrated in females based on diagnostic history. In this scenario, each step-up in risk associated with the given diagnoses is associated with a steeper increase in risk. The greater associations observed among females may be related to gendered differences in substance use behaviour—for instance, women with depression may use substances alone, which can increase overdose risk ^55^. The decision to use substance alone may also reflect macro/structural and interpersonal factors: for instance, people with lived and living experience of substance use have reported that women may alter substance use behaviour, including by using alone, due to the risk of physical and sexual violence associated with the sedating effects of the unregulated drug supply ^56,57^.

When comparing overall IRRs to the male-stratified IRRs, the depression effect was slightly smaller in the overall estimates than in males alone, indicating confounding by sex, consistent with the higher prevalence of documented depression among females, who have lower overdose incidence. Additionally, overall results showed an attenuation of effect sizes when comparing NFOD to FOD; stratifying by sex indicates this is mostly due to the trends for females, and the same does not apply for males. Taken together, these points suggest that analyses not considering sex and/or gender may underestimate the association between documented diagnosis history and overdose risk for both males and females

The associations between depression and SUD history and SUD history and overdose risk were extremely strong. This is somewhat to be expected given prior research indicating that a substantial proportion of OD cases in the province have a documented SUD diagnosis^39^ despite a very small proportion of the BC population having a documented SUD diagnosis ^53^. Differences in the age distribution of the Depression and SUD and SUD versus the depression and neither groups may be associated with the higher all-cause mortality among people with SUD ^58,59^.

The high rates of recurrent and prior NFOD observed across all diagnostic groups, appearing most elevated among those with documented depression and SUD and SUD histories, followed by those with a history of depression diagnosis alone, and then those with neither diagnosis, suggest that a first NFOD represents an important intervention point. The elevated recurrence risk among those with a documented depression diagnosis history without SUD may also indicate that this is true not only for SUD-focused care, but also for integrating mental health and depression care as well.

The contribution of this analysis to the literature is that it directly isolates the effects of a documented depression diagnosis history from those of a documented SUD history, describing how a documented depression diagnosis operates as its own risk factor, and how this may differ by sex. This may be of particular interest given the stronger association between documented depression history and overdose risk in the 19-29 group for females, and the context of surveillance data from the US linking mental health concerns among female youth to increases in youth overdose rates ^2,60^. There is also a growing body of literature from Canada, in addition to the US and some European countries, indicating disproportionately worsening mental health among female youth ^6,61,62^, underscoring the timeliness and relevance of this study.

The age- and sex-patterned findings may also be relevant for older age groups, where recent surveillance data in BC indicate that FOD rates are not declining among those above 60 years of age, despite declines in other age groups ^1^. Notably, this study’s findings suggest that the association between a documented depression, SUD, or Depression and SUD diagnosis and overdose risk increases with age, potentially peaking at the 60-69 age group. As well, a documented SUD history alone overtakes depression and SUD history in terms of effect size in older age groups among both males and females, though confidence intervals overlapped and differences were modest, suggesting an area for potential monitoring and future research. This could also be related to differences in depression and SUD risk over the life course ^63,64^, alongside sex and gender differences in the mental health of older adults and social isolation in this population ^65^. Additionally, age-period-cohort analyses of overdose in BC suggest that increases in overdose rates were greatest among Gen X and Baby Boomer males during the pandemic period, followed by the greatest subsequent decreases in these groups ^66^. Taken together with this study’s findings, it appears that there may be a pattern involving age, sex, and mental health diagnosis history that warrants further and more targeted investigation to identify interventions to reduce overdose in this population.

### Strengths

This study used a population-based sample with data linked across multiple comprehensive datasets, providing broad coverage and reducing selection bias. The extended follow-up period captured multiple phases of the unregulated drug poisoning emergency, including the initial rise following the declaration of an emergency, increases during and following the COVID-19 pandemic, and the beginnings of a decline in fatalities starting in 2024^1^, allows this study to provide a more robust description of the association between the documented diagnosis histories in question and overdose risk than studies using a more restricted time period. Finally, this study used case definitions and algorithms used for surveillance and decision making in the province, strengthening the applicability of the findings across policy contexts.

### Limitations

Several limitations of this study should be noted. The NFOD definition used in this study has linkage limitations; specifically, prior analyses of 137,893 overdose calls recorded between January 1^st^, 2015, to December 31^st^, 2023, identified 28,528 that could not be linked due to limited personal identifiers ^67^. These unlinkable events differed from linkable events, such as by being more likely to occur in the Vancouver Coastal Health region or on roadways, and thus their exclusion may introduce bias (R. Desai, Unpublished data, 2026). Further, the people whose healthcare contacts more broadly cannot be linked using the CLR, as described in Section 2.2.3, may be more likely to be structurally and/or socially marginalized ^34^, which could, given the relationships between overdose, mental health, and marginalization, lead to underestimates of the associations between diagnosis history and overdose risk^68,69^.

The use of a binary sex variable, as recorded in administrative data, also carries several limitations. Shifts in the ability to change sex markers on government identification mean that sex markers in administrative health data may inconsistently reflect gender identity for some people depending on whether a person has updated their identification or not, limiting the precision of this variable. More broadly, a binary sex variable cannot capture trans and gender-diverse identities, who may be at higher risk of depression and/or SUD diagnoses ^70^. Additionally, using a solely binary sex variable may limit the ability to distinguish which of the observed effects are due to biological or sex-based factors, and which are due to socio-cultural and gender-based factors.

With respect to the diagnostic exposure groups, documented depression and SUD diagnosis histories were ascertained within a defined lookback window, so some of the cases identified may not be an active diagnosis case at the time of overdose, while conversely some “neither” cases have an undetected diagnosis. Specifically, some individuals in the “neither group” received a documented SUD, depression, or combined diagnosis between the end of the ascertainment window and their NFOD or FOD event (see Appendices A.3 and A.4). This was most common among those who experienced a FOD. As a result, the strength of the associations between different diagnostic histories and overdose risk reported in this study may be underestimates. Further, these diagnoses only reflect cases among those who sought care in a manner that could be captured within the health administrative data used, and thus these groups are more indicative of peoples’ healthcare history, rather than a definitive classification. Finally, ICD code 50B, which is included in the depression case definition, captures both anxiety and depression, meaning some anxiety cases may be misclassified as depression cases.

The estimates in this study were largely descriptive in nature and are not adjusted for other factors that influence overdose risk, such as material and social deprivation, rurality, regional health authority, and other comorbid health conditions ^39,71^. Of particular interest, some of the findings regarding age by sex differences may be explained in part by ethnicity and Indigeneity, given the disproportionately high OD rates among female Indigenous youth ^72^. Other factors not included in the analysis such as material and social deprivation, region of residence, and housing status, which were differentially distributed among those who experienced a NFOD or FOD, and could also be contributing to some of the observed sex differences in the associations between depression and/or SUD history and overdose risk (see Appendix A.5)^73–75^. Lower overall case counts for OD for females may affect the reliability of the estimates for females, particularly for the age-stratified estimates. The higher incidence of OD in the “neither” group among the 19-29 age group, and the relative underrepresentation of this age group in the different diagnostic categories, may be in part due to differences in when these conditions tend to be diagnosed.

Finally, some of the FOD and NFOD events in the “neither” group among males may be attributable to severe mental illnesses that are more common among males and increase OD risk^20,76^ but were not measured in this study.

## Data Availability

Requests for access to the data in the present study can be made directly through correspondence with the corresponding author

## Acknowledgements

- Access to data provided by the Data Stewards is subject to approval, but can be requested for research projects through the Data Stewards or their designated service providers. All inferences, opinions, and conclusions drawn in this publication are those of the author(s), and do not reflect the opinion of the Data Stewards. The following data sets were used in this project: Medical Services Plan (MSP), Discharge Abstract Database (DAD), BC Emergency Health Services (BCEHS), BC Coroners Service (BCCS), and Client Roster (CLR), National Ambulatory Care Reporting System (NACRS). This Data was provisioned under ISP 21-051.
- I gratefully acknowledge the residents of British Columbia whose data are integrated in the data sets we used on PANDA.
- I acknowledge the Professionals for the Ethical Engagement of Peers (PEEP) and First Nations Health Authority (FNHA) Health Surveillance team for their feedback on this work.

## Appendices

**A.1.**
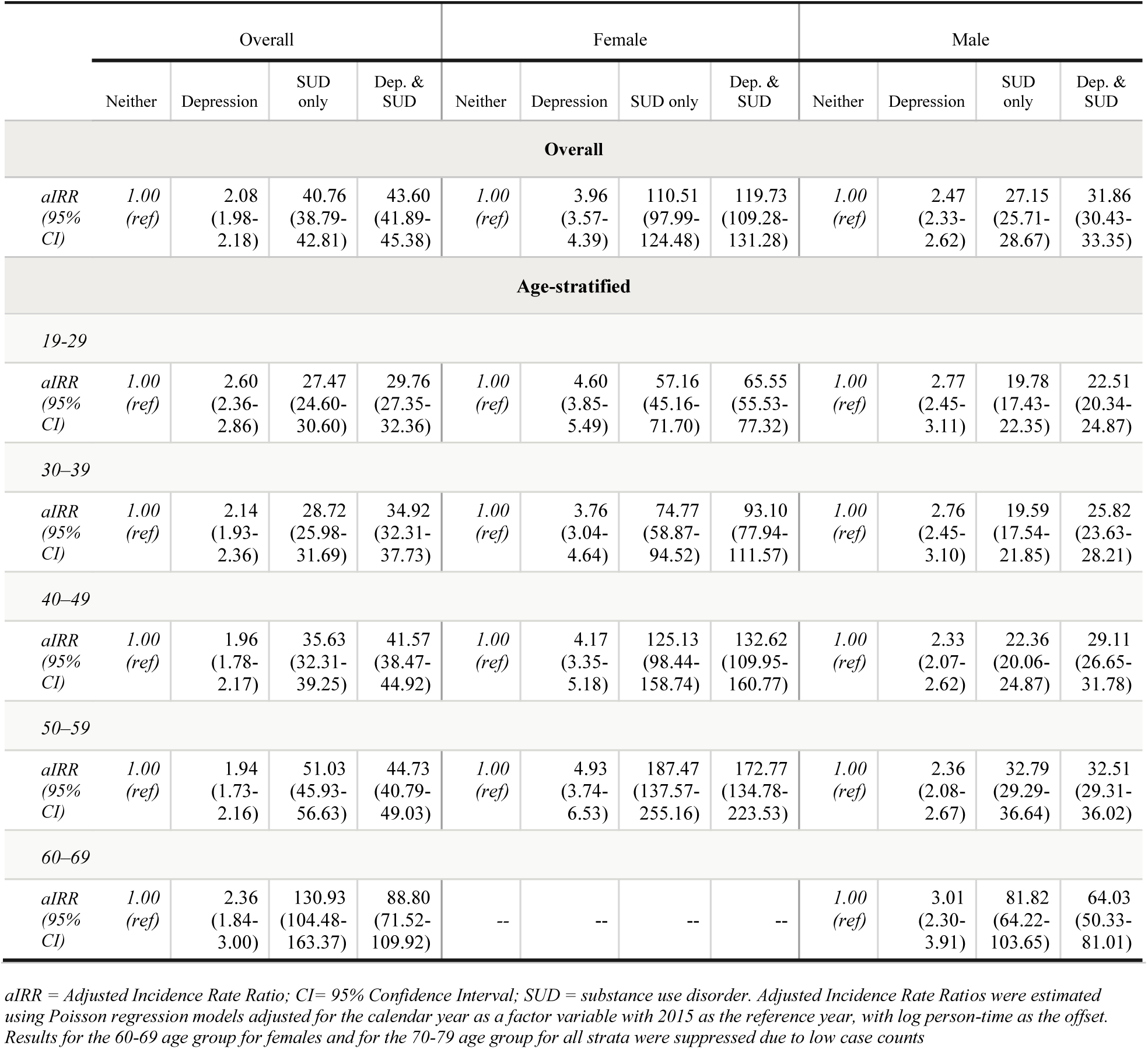
Year-Adjusted Incidence Rate Ratios for Fatal Overdose by Diagnostic History Group, Overall and Stratified by Sex and Age Group, British Columbia, 2015-2024.

**A.2.**
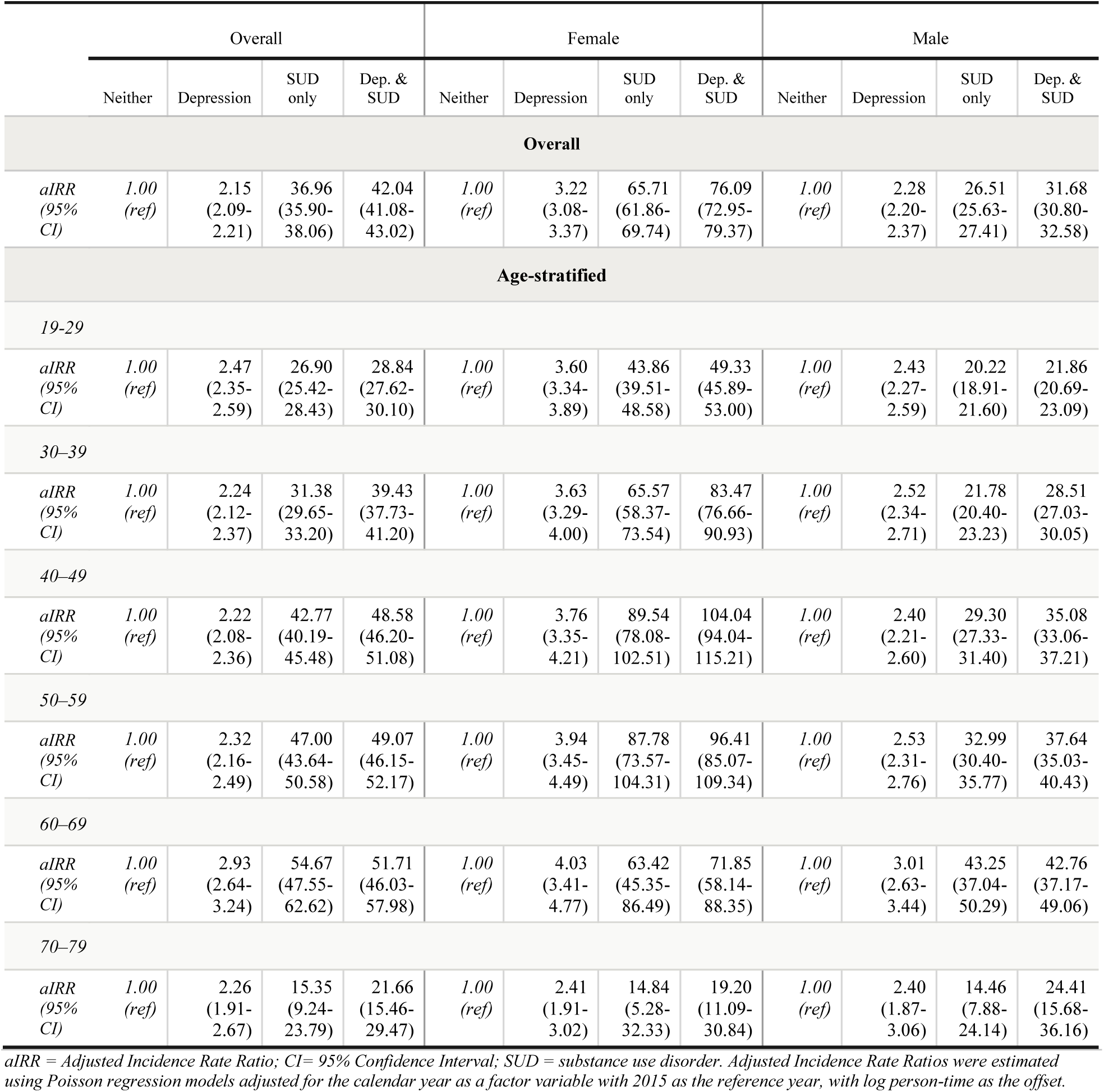
Year-adjusted Incidence Rate Ratios for Non-Fatal Overdose by Diagnostic History Group, Overall and Stratified by Sex and Age Group, British Columbia, 2015-2024.

**A.3.** Subsequent Mental Health and Substance Use Diagnoses Among Cohort Members with No Baseline Diagnosis ("Neither") who Experienced a Non-Fatal Overdose.

| Diagnosis received during follow-up | Neither — NFOD<br>*N* = 18,936<br>*N* (%) |
| --- | --- |
| Depression only | 2,032 (10.73) |
| SUD only | 2,921 (15.43) |
| Depression and SUD | 2,355 (12.43) |
| No subsequent diagnosis | 11,628 (61.41) |

SUD = substance use disorder. NFOD = Non-fatal overdose.
"Neither" category defined as cohort members with no recorded depression or SUD diagnosis in between January 1, 2010 and December 31, 2014. Subsequent diagnoses were identified between January 1, 2015 and the date of the person's fatal overdose.

**A.4.** Subsequent Mental Health and Substance Use Diagnoses Among Cohort Members with No Baseline Diagnosis ("Neither") who Experienced a Fatal Overdose.

| Diagnosis received during follow-up | Neither — FOD<br>*N* = 5,979<br>*N* (%) |
| --- | --- |
| Depression only | 587 (9.82) |
| SUD only | 1,175 (19.65) |
| Depression and SUD | 1,015 (16.98) |
| No subsequent diagnosis | 3,202 (53.55) |

SUD = substance use disorder. FOD = Fatal overdose.
"Neither" category defined as cohort members with no recorded depression or SUD diagnosis in between January 1, 2010 and December 31, 2014. Subsequent diagnoses were identified between January 1, 2015 and the date of the person's non-fatal overdose.

**A5.** Additional Demographic Characteristics of the Full Study Cohort and Those who Experienced a Fatal or Non-Fatal Overdose in British Columbia, 2015-2024.

|  | Entire Study Cohort<br>*N* = 3,996,690<br>*N* (%) | Fatal Overdose<br>*N* = 14,281<br>*N* (%) | Non-Fatal Overdose<br>*N* = 43,555<br>*N* (%) |
| --- | --- | --- | --- |
| **HEALTH AUTHORITY** |  |  |  |
| N/A | 104,314 (2.61) | 130 (0.91) | 585 (1.34) |
| Fraser | 1,029,524 (25.76) | 2,787 (19.52) | 7,772 (17.84) |
| Interior | 477,666 (11.95) | 1,109 (7.77) | 3,348 (7.69) |
| Northern | 322,837 (8.08) | 2,081 (14.57) | 6,820 (15.66) |
| Vancouver Coastal | 1,272,938 (31.85) | 4,583 (32.09) | 13,382 (30.72) |
| Vancouver Island | 789,411 (19.75) | 3,591 (25.15) | 11,648 (26.74) |
| **MATERIAL DEPRIVATION (QUINTILE)** |  |  |  |
| N/A | 60,857 (1.52) | 15 (0.11) | 25 (0.06) |
| Q1 (least deprived) | 875,792 (21.91) | 2,695 (18.87) | 5,271 (12.10) |
| Q2 | 818,391 (20.48) | 2,134 (14.94) | 5,878 (13.50) |
| Q3 | 795,881 (19.91) | 2,628 (18.40) | 6,595 (15.14) |
| Q4 | 779,836 (19.51) | 3,145 (22.02) | 9,247 (21.23) |
| Q5 (most deprived) | 665,933 (16.66) | 3,664 (25.66) | 16,539 (37.97) |
| **SOCIAL DEPRIVATION (QUINTILE)** |  |  |  |
| N/A | 60,857 (1.52) | 15 (0.11) | 25 (0.06) |
| Q1 (least deprived) | 742,311 (18.57) | 1,635 (11.45) | 7,614 (17.48) |
| Q2 | 749,104 (18.74) | 1,798 (12.59) | 6,890 (15.82) |
| Q3 | 734,193 (18.37) | 2,013 (14.10) | 8,323 (19.11) |
| Q4 | 834,001 (20.87) | 2,939 (20.58) | 9,686 (22.24) |
| Q5 (most deprived) | 876,224 (21.92) | 5,881 (41.18) | 11,017 (25.29) |
| **NO FIXED ADDRESS** |  |  |  |
| No | 3,978,908 (99.56) | 12,565 (87.98) | 38,877 (89.26) |
| Yes | 17,782 (0.44) | 1,716 (12.02) | 4,678 (10.74) |

Fatal overdose was ascertained from BC Coroners Service Records; non-fatal overdose was ascertained from BC Emergency Health Services paramedic-attended overdose records. Individuals may appear in both the FOD and NFOD columns if they experienced both outcomes during follow-up. Material and Social Deprivation Quintiles were obtained from INSPQ indices. No fixed address refers to history of a health record indicating no fixed address between 2010-2014, as indicated by DAD and NACRS records.

